# Pain, inconvenience, and blame - Defining work-related injuries in the veterinary workplace

**DOI:** 10.1101/2023.12.14.23299902

**Authors:** Tamzin Furtado, Martin Whiting, Imogen Schofield, Rebecca Jackson, John S.P. Tulloch

## Abstract

**Objectives:** The veterinary workplace carries a high risk of staff accidents and injuries, yet there is scant research exploring it in comparison with other comparable fields, such as human medicine. The aim of this study was to understand how veterinary professionals define injuries and to understand what injuries they do, or do not, deem reportable.

**Methods:** A cross-sectional survey comprising demographic questions and open-text questions was shared with veterinary practice staff across the United Kingdom. Data were analysed descriptively and using an inductive content analysis.

**Results:** There were 740 respondents, who were broadly representative of the veterinary profession. There were differences in how injuries were defined; for example, small animal veterinarians expected injuries to involve blood, while equine and production animal veterinarians were more likely to expect injuries to reduce their ability to perform work and result in time off work. Many suggested that “all” workplace injuries should be reported, however “minor” injuries were often overlooked, for example needlestick injuries did not always meet the criteria of being an “injury”. Injuries caused by staff themselves (e.g. trips) were less likely to be reported than injuries that could be blamed on an external factor (e.g. dog bite).

**Conclusions:** Collectively, the data suggest a wide-ranging perception of risk of injury in practice, with some harms seen as “everyday norms”. Veterinary practices should interpret their injury statistics with a high degree of caution. They should explore the microcultures within their practices relating to worker perception of risk, injury and barriers to reporting.

**What is already known on this topic:** The veterinary industry has one of the highest case rates of non-fatal occupation injuries and illnesses per full time worker. In the USA, no other industry is higher; it is almost five times higher than the national average. Yet, little research has explored how injuries are perceived nor their context.

**What this study adds:** This study shows clear divisions within different veterinary sectors, and job roles, in how injuries are perceived. In particular, that equine and production animal veterinarians have a high threshold before acknowledging that an incident is a work-related injury.

**How this study might affect research, practice or policy:** To contextualise any epidemiological research into veterinary workplace injuries, one needs to understand how injuries are perceived. The discordance in definition needs to be accounted for when interpreting company or national injury reporting figures.

## Introduction

The veterinary industry has one of the highest rates of work-related injuries. In the United States of America (USA) it has the highest levels of non-fatal occupation injuries and illness (13.8 cases per 100 full time workers in 2021), almost five times higher than the national average (1,2). In the United Kingdom (UK), being an equine veterinarian has been deemed the most dangerous civilian (3). Potential hazards include; the physical working environment, machinery, sharp equipment, toxic materials, medications, and animals (4,5). However, veterinary workplaces have received little attention in relation to the sociological and epidemiological study of such injuries, in comparison with other medical settings.

While workplace safety has generally improved, there are still a relatively high number of workplace injuries across some industries, and a relatively poor understanding of the psychology and human factors which contribute to them (6). Some suggest that the focus on reporting systems has increased scepticism around health and safety management, while doing little to improve actual outcomes for people in workplaces (7).

Exploration of work-related injuries must start by considering how industry specific employees define a work-related injury. The Centers for Disease Control and Prevention (CDC) define an injury as; ‘*A bodily harm resulting from severe exposure to an external force or substance (mechanical, thermal, electrical, chemical, or radiant) or a submersion. This bodily harm can be unintentional or violence-related*’ (8). The Health and Safety Executive (HSE) state that for an injury to be work-related it ‘*must contribute to the accident. An accident is ‘work-related’ if any of the following played a significant role: the way the work was carried out; any machinery, plant, substances or equipment used for the work or, the condition of the site or premises where the accident happened*’ (9). Accident reporting systems rely on individuals being able to identify when a work-related accident has occurred and then report it. If an employee utilises a different work-related injury definition, then reporting rates will not be reflective of the true extent of injuries.

Differing job roles in an industry may perceive different levels of acceptable risk (10), and how they define an injury (11). Additionally, emotional intelligence is associated with lower levels of injury, suggesting that psychological and emotional traits, such as calmness, might impact perception of safe practices, as well as behaviour (12). Different workplace activities, professional values, and work cultures will exist within any given industry and workplace, and these will also contribute to what people perceive to be an injury and thus how people recognise and respond to work-related risk.

The reasons for injury underreporting include: a lack of trust that change will occur; lack of time; lack of familiarity with reporting practices; concern over getting other staff members in trouble; and concern regarding blame(13,14). Veterinary work is known to be both highly risky and highly specific in nature, therefore we might expect that veterinary perception of accident reporting to differ from other industries. However, beyond establishing that veterinary work presents a high level of risk, particularly for production animal and equine veterinarians (2–4), the veterinary industry has been predominantly overlooked.

This study aims to understand how the veterinary professions define injuries and what type of injury they do, or do not, deem reportable. This information can be used to support veterinary practices in exploring their own approaches to risk and accident identification and reporting, and thus in creating safer workspaces.

## Methods

An online questionnaire was developed, comprising a mix of categorical, numerical, and open-ended questions. Participants were asked about personal demographics (i.e. sex, age) and their work (i.e. the type of practice, their job role). They were asked to provide their own definition of a work-related injury, to describe a work-related injury that they felt needed reporting, and to describe a work-related injury that did not need reporting. The questionnaire was piloted with a sample of veterinary professionals.

The survey was distributed to all UK employees of CVS UK Ltd (CVS) which comprises approximately 500 veterinary practices. The survey was distributed through an internal newsletter and was open for three months between 6^th^ December 2022 and 6^th^ March 2023. To enhance recruitment, an incentive of a year’s supply of snacks for five randomly selected practices was provided.

All responses were stratified by job role. Descriptive statistics were performed to describe the respondent population, and open-ended questions were analysed using inductive qualitative content analysis (15,16), which provides a flexible approach combining elements of qualitative analysis with quantitative components. Each response was initially read through, whilst notes describing content categories were created, then each response was considered in isolation and was codified to reflect the information within that item. Codes were created in an iterative fashion, and as more codes were created, they were combined, revised, and deleted. The final set of codes were generated by repetition of this process to ensure internal validity of content in the codes across the dataset. Instances of each code occurring were then counted to facilitate a quantitative comparison of content.

Data masking was used to protect personally identifiable information of respondents.

## Results

Over a thousand individuals (n=1102) consented to the survey, with 740 (67.2%) responses that were complete enough for analysis. Of the responses, veterinarians were the most prevalent job role (250; 33.8%), of whom 74.8% were companion animal veterinarians. Other roles included veterinary nurses (28.1%), administrators (15.5%), receptionists (14.3%), and companion animal Patient Care Assistants (PCA) (8.2%) (Table 1).

**Table 1.** Veterinary work-related injury survey respondent demographics.

|  | Equine Veterinarian (n=47) | Production Animal Veterinarian (n=16) | Companion Animal Veterinarian (n=187) | Total Veterinarians (n=250) | Companion Animal Veterinary Nurse (n=208) | Companion Animal Patient Care Assistant (n=61) | Administrator (n=115) | Receptionist (n=106) | Total (n=740) |
| --- | --- | --- | --- | --- | --- | --- | --- | --- | --- |
| **Sex (% Female)** | 59.6% (44.3-73.6) | 81.3% (54.4-96.0) | 83.4% (77.3-88.5) | 78.8% (73.2-83.7) | 94.2% (90.1-97.0) | 86.9% (75.8-94.2) | 91.3% (84.6-95.8) | 99.1% (94.9-100.0) | 88.6% (86.1-90.8) |
| **Age** |  |  |  |  |  |  |  |  |  |
| **<20** | 0 | 0 | 0 | 0 | 0 | 1.6% (0.0-8.8) | 0 | 1.9% (0.2-6.7) | 0.4% (0.1-1.2) |
| **20-29** | 14.9% (6.2-28.3) | 37.5% (15.2-64.6) | 23.0% (17.2-29.7) | 22.4% (17.4-28.1) | 28.8% (22.8-35.5) | 54.1% (40.9-66.9) | 9.6% (4.9-16.5) | 33.0% (24.2-42.8) | 26.4% (23.2-29.7) |
| **30-39** | 36.2% (22.7-51.5) | 37.5% (15.2-64.6) | 36.9% (30.0-44.3) | 36.8% (30.8-43.1) | 35.1% (28.6-42.0) | 26.2% (15.8-39.1) | 20.0% (13.1-28.5) | 14.2% (8.1-22.3) | 29.6% (26.3-33.0) |
| **40-49** | 27.7% (15.6-42.6) | 18.8% (4.0-45.7) | 25.7% (19.6-32.6) | 25.6% (20.3-31.5) | 28.8% (22.8-35.5) | 6.6% (1.8-16.0) | 41.7% (32.6-51.3) | 12.2% (6.7-20.1) | 25.5% (22.4-28.8) |
| **50-59** | 14.9% (6.2-28.3) | 6.3% (0.2-30.2) | 9.6% (5.8-14.8) | 10.4% (6.9-14.9) | 5.3% (2.7-9.3) | 8.2% (2.7-18.1) | 21.7% (14.6-30.4) | 21.7% (14.3-30.8) | 12.2% (9.9-14.7) |
| **60+** | 6.4% (1.3-17.5) | 0 | 4.8% (2.2-8.9) | 4.8% (2.5-8.2) | 1.9% (0.5-4.9) | 3.3% (0.4-11.4) | 7.0% (3.1-13.3) | 17.0% (10.4-25.5) | 5.9% (4.4-7.9) |
| **Ethnicity** | (n=46) |  | N=186 | 248 | 207 |  | 113 |  | 735 |
| **White** | 97.8% (88.5-100.0) | 100.0% | 95.2% (91.0-97.8) | 96.0% (92.7-98.1) | 97.1% (93.8-98.9) | 98.4% (91.2-100.0) | 98.2% (93.8-99.8) | 98.1% (93.4-99.8) | 97.1% (95.7-98.2) |
| **All other ethnic groups combined** | 2.2% (0.1-11.5) | 0 | 4.8% (2.2-9.0) | 4.0% (2.0-7.3) | 2.9% (1.1-6.2) | 1.6% (0.0-8.8) | 1.8% (0.2-6.2) | 1.9% (0.2-6.7) | 3.0% (1.9-4.5) |
| **Ever had an injury** | 97.9% (88.7-100.0) | 93.8% (69.8-99.8) | 93.6% (89.1-96.6) | 94.4% (90.8-96.9) | 92.8% (88.4-95.6) | 75.4% (62.7-85.5) | 50.4% (41.0-59.9) | 62.3% (52.3-71.5) | 80.9% (77.9-83.7) |

All respondents who provided their sex, also identified with the same gender. Veterinarian respondents were representative of the UK profession in terms of age (median national age 30-39) and ethnicity (3.5% ethnic minority groups nationally) (17). However, female respondents were over-represented (59% of veterinarians are female nationally). Companion animal veterinary nurses were broadly representative in terms of sex (96.8% female nationally), age (median age of 30-39 nationally), and ethnicity (1.9% ethnic minority groups nationally) (18). National demographics of companion animal PCAs, administrators, and receptionist are currently not available.

The prevalence of work-related injuries experienced during their career was the highest for veterinarians (94.4%), whilst the lowest were administrators (50.4%). Of the veterinarians, the highest injury prevalence was seen in the equine veterinarian group, where 97.9% had experienced an injury.

### 1. How do you define an injury?

When asked to define an injury, participants most commonly referred to elements of physical trauma (Table 2). Descriptions coded as “physical trauma” included references to physical injury, damage to the body, or harm:

**Table 2.**
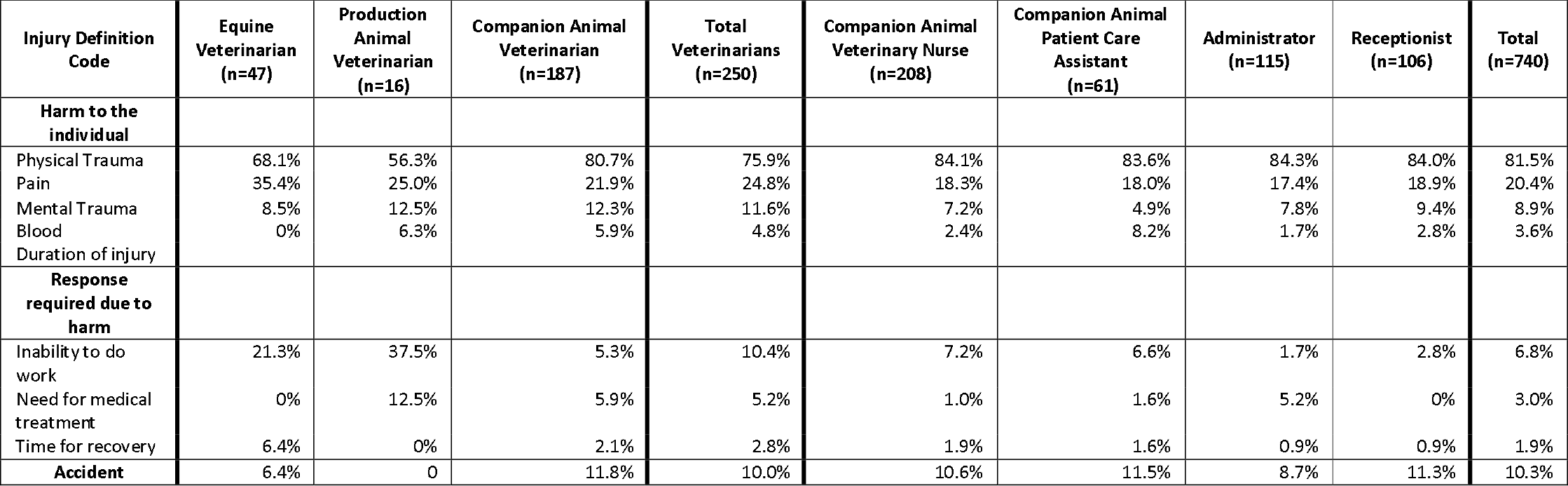
Content analysis of the veterinary profession’s definition of an injury.

> “*Any incident involving harm to a person*” (companion animal veterinary nurse)
>
> “*Any damage to the body*” (companion animal veterinarian)

Many participants provided greater depth to what they would consider a physical trauma in relation to a workplace accident, and these codes were divided into three themes *harm to the individual* (codes included: blood, pain, or mental harm), *response required due to harm* (codes included: need for medication/treatment, time for recover, inability to perform job role), and *accident*. In terms of *harm to the individual*, many participants explicitly stated they would consider something an injury if there was a high degree of severity:

> “*An incident that results in a cut/bruise/wound/fracture/dislocation/pain or damage to my body*” (companion animal veterinarian)
>
> “*An occurrence that causes pain or visible signs (i*.*e back pain, broken skin, bruising)*” (companion animal administrator)

Some participants described that pain was a core component of how they perceived an injury. The code for “pain” was used more by equine and production animal veterinarians than other roles (35.4% and 25%, compared with 21.9% for companion animal veterinarians), and tended to be used as a descriptive additive clause to physical harm.

> *“An injury is usually an event that happens where your body is hurt and elicits excessive pain/inconvenience”* (production animal veterinarian)

Mental trauma was referenced by many, though always in addition to physical harm:

> “*An event which causes tissue or psychological damage*” (companion animal veterinarian)

Production animal and companion animal veterinarians tended to use “mental trauma” more than other roles (12.5% and 12.3% respectively, compared with 8.5% for equine veterinarians and 7.2% for veterinary nurses). None of the participants gave more detail about the extent or duration of mental trauma that they felt constituted an “injury”, which contrasted with the physical trauma definitions.

In relation to the theme responses required due to harm; many participants described that a core component of their definition of “injury” was the need to take action, for example to seek medical treatment, take time to recover, or alter their behaviour:

> *“Something that requires me to open the first aid box or ‘take a minute’ to recover from while doing my job”* (companion animal veterinarian)
>
> *“Where you have hurt yourself and need to go to hospital or it leaves a mark or pain longer than 1 day*.*”* (production animal veterinarian)

Notably, this ‘need for medical treatment’ was most prevalent for production animal veterinarians, at 12.5% of respondents, compared with 0% of equine veterinarians and 5.9% of companion animal veterinarians.

The code ‘inability to work’ within this theme had the largest difference in prevalence between job roles. For equine and production animal veterinarians it was used in 21% and 38% of definitions, compared to less than 10% in all other veterinary roles. It was primarily used in isolation;

> *‘Any injury that prevents me carrying out my role as a practising veterinary surgeon*.*’* (production animal veterinarian)

Contrastingly, other job roles tended to combine it with other themes, such as pain:

> *‘Hurt / pain with tissue damage and reduced capacity to work to full potential*.*’* (companion animal veterinarian)

Overall, the theme of ‘accident’ was the third most prevalent. This was rarely used in isolation but as a conjunctive to ‘physical trauma’:

> *“Accidental trauma, hurt or damage. physical or mental*.” (companion animal veterinarian)
>
> *“An injury is something that has caused harm to your body through an accident*.*”* (companion animal administrator)

This suggests that these participants considered accidental harm differently to harms caused by, for example, negligence.

### 2 What type of injuries need reporting?

When asked to describe a work-related injury that they thought needed reporting to their employer, a large proportion (39.5%) of individuals stated that all work-related injuries should be recorded (Table 3), for example:

**Table 3.** Content analysis of the veterinary profession’s definition of a work-related injury that they would report to an employer.

| Reportable injury code | Equine Veterinarian (n=47) | Production Animal Veterinarian (n=16) | Companion Animal Veterinarian (n=187) | Total Veterinarians (n=250) | Companion Animal Veterinary Nurse (n=208) | Companion Animal Patient Care Assistant (n=61) | Administrator (n=115) | Receptionist (n=106) | Total (n=740) |
| --- | --- | --- | --- | --- | --- | --- | --- | --- | --- |
| **All** | 40.4% | 43.8% | 33.7% | 35.6% | 38.5% | 34.4% | 47.0% | 45.2% | 39.5% |
| **None** | 0% | 0% | 0.5% | 0.4% | 0% | 1.6% | 0.9% | 1.9% | 0.7% |
| **Modifier** |  |  |  |  |  |  |  |  |  |
| Severity | 27.7% | 31.3% | 15.0% | 18.4% | 5.3% | 1.6% | 4.3% | 8.5% | 9.7% |
| Impacted ability to work | 10.6% | 25.0% | 7.0% | 8.8% | 2.9% | 6.6% | 5.2% | 1.9% | 5.4% |
| Medical Treatment | 6.4% | 6.3% | 3.2% | 4.0% | 2.9% | 1.6% | 5.2% | 2.8% | 3.5% |
| **Examples** |  |  |  |  |  |  |  |  |  |
| Bite or Scratch | 0% | 0% | 46.0% | 34.4% | 42.3% | 34.4% | 20.0% | 14.2% | 31.5% |
| Kick or trample | 36.2% | 12.5% | 0% | 7.6% | 0% | 0% | 5.2% | 0.9% | 3.5% |
| Slip or fall | 0% | 0% | 5.3% | 4.0% | 7.2% | 8.2% | 9.6% | 21.7% | 8.6% |
| Needlestick | 0% | 6.3% | 9.1% | 7.2% | 6.3% | 6.6% | 3.5% | 3.8% | 5.8% |
| Equipment | 4.3% | 6.3% | 1.6% | 2.4% | 3.8% | 1.6% | 9.6% | 1.9% | 3.9% |
| Musculoskeletal injuries | 2.1% | 6.3% | 2.1% | 2.4% | 1.9% | 1.6% | 0% | 5.7% | 2.4% |
| Burn | 0% | 0% | 0% | 0% | 1.0% | 4.9% | 0% | 1.9% | 0.9% |

> “*Any injury sustained in the workplace*” (companion animal veterinarian)
>
> “*Any injury at all including near misses*” (companion animal veterinarian)

Many provided detail which modified their answer, for example stating that while all injuries should be reported, they would only report an injury if: it was severe; it impacted their ability to work; or it required medical treatment. Overall, the use of these modifiers was more prevalent in veterinarians than other roles, and in particular equine and production animal veterinarians. For example:

> *“I think you’re supposed to report all of them, but I tend to report only ones that impact my ability to keep working or need intervention to keep working*” (companion animal veterinarian)
>
> *“Technically any injury should be reported especially those related to poor maintenance or set up. I think any injury that may result in seeing a Dr or any time off should be reported*.*”* (equine veterinarian)

These responses highlight the gap between the ways people think they should behave, and how they actually behave, which involved individual estimation of the severity of the injury or requirement of additional support. One participant described her reasons for not reporting in more detail:

> *“I’ve perhaps reported some of them but the pace of work is so fast and as I feel it’s not going to change anything for me then I report very few of them*.*”* (companion animal veterinarian)

When respondents gave examples of injuries that needed reporting, the majority described injuries caused by animals (i.e. bites, kicks), or caused by hazards (i.e. slips, trips, and falls). These injuries could be positioned as being caused by other parties, such as the environment, humans or animals:

> *“A fall that was at the result of something in the practice*” (companion animal veterinary nurse)

> “*Common injuries in our profession are often bite related injuries from handling animals*” (companion animal veterinary nurse)

When asked to describe a work-related injury they thought did not need reporting, a large proportion of respondents said that they would report all injuries. This was lowest amongst equine and production animal veterinarians who were more likely to provide specific examples of cases that would not need reporting (Table 4).

**Table 4.** Content analysis of the veterinary profession’s definition of a work-related injury that they would not report to an employer.

|  | Equine Veterinarian (n=47) | Production Animal Veterinarian (n=16) | Companion Animal Veterinarian (n=187) | Total Veterinarians (n=250) | Companion Animal Veterinary Nurse (n=208) | Companion Animal Patient Care Assistant (n=61) | Administrator (n=115) | Receptionist (n=106) | Total (n=740) |
| --- | --- | --- | --- | --- | --- | --- | --- | --- | --- |
| **None** | 19.1% | 18.8% | 26.7% | 24.8% | 38.0% | 34.4% | 63.5% | 42.4% | 37.8% |
| **Modifier** |  |  |  |  |  |  |  |  |  |
| Minor | 42.5% | 31.3% | 21.9% | 26.4% | 14.9% | 19.7% | 7.0% | 16.0% | 18.1% |
| Self-attribution | 8.5% | 18.8% | 8.0% | 8.8% | 12.0% | 3.3% | 4.3% | 6.6% | 8.2% |
| No Medical treatment | 12.8% | 0% | 4.3% | 5.6% | 3.4% | 3.3% | 0.9% | 0.9% | 3.4% |
| No blood | 0% | 0% | 5.3% | 4.0% | 0.5% | 1.6% | 0% | 0.9% | 1.8% |
| **Example** |  |  |  |  |  |  |  |  |  |
| Papercut | 8.5% | 6.3% | 11.2% | 10.4% | 15.3% | 18.0% | 14.8% | 27.4% | 15.5% |
| Bite and Scratch | 0% | 0% | 15.0% | 11.2% | 7.2% | 14.8% | 0.9% | 2.8% | 7.6% |
| Needlestick | 4.3% | 25.0% | 7.5% | 8.0% | 5.3% | 0% | 1.8% | 0.9% | 4.6% |
| Fall or Slip | 0% | 0% | 0.5% | 0.4% | 1.4% | 3.3% | 1.8% | 1.9% | 1.4% |
| Kick | 14.9% | 6.3% | 0% | 3.2% | 0.5% | 0% | 0% | 0% | 1.2% |

Veterinarians, in particular, would not report if they felt the injury was minor or if they required no medical treatment; equine veterinarians were the least likely to report injuries which they considered minor (42.5% would not report minor incidents, compared with 26.4% for veterinarians overall).

> “*Something that does not require First Aid is not an injury that needs to reporting*.” (companion animal care assistant)

Notably, 8.0% of veterinarians gave a needlestick injury as an example of an injury that did not need reporting:

> *“every day needle stick injury, occurs quite commonly, as long as not a dangerous drug and minor injury”* (companion animal veterinarian)

Companion animal veterinarians were more likely than other veterinarians to mention a lack of blood as a reason for not reporting (5.3% of small animals mentioned lack of blood, compared with no equine or production animal vets). This is likely to relate to common injury types experienced, as 15% of small animal veterinarians also used bites or scratches as examples of minor injuries, compared with 14.9% of equine vets who used a kick as an example of an injury that did not need reporting:

> “*Vet knocked into wall by horse causing pain initially but no visible bruises or residual pain*” (equine veterinarian)
>
> “*In this job role we probably get scratched quite a lot by animals. Unless its a really bad/deep scratch I wouldn’t report it to my employer*.” (companion animal veterinary nurse)

Participants frequently described self-assessing the severity of injury and the requirement for behaviour change to avoid future instances, when deciding whether to report an injury.

> *“Where the injury was deemed insignificant by the person/people involved and where people feel confident that procedures/protocols don’t need to be changed, or awareness raised, to prevent future problems*.*”* (companion animal veterinarian)

This self-assessment process could include an estimation of whether the accident could have been avoidable, for example by following protocols or having a clear idea of health and safety procedures.

> *“Any injury that you feel is caused by your own lack of judgement/ignorance of health and safety rules and that does not require any treatment or time off work*” (companion animal veterinarian)

This response also displays an additional theme: self-attribution of injury cause. In contrast to the previous question where reported items were often the result of interaction with an animal or working environment which caused harm, it was notable in this item that participants considered injuries which did not need reporting to be those which they felt were ‘self-inflicted’. For example, a papercut was the most frequently cited injury that was perceived as not needing reporting across all groups; other examples included:

> *“Hurting yourself by being stupid i*.*e. walking into a door”* (companion animal veterinary nurse)
>
> *“bruising leg by walking into table”* (companion animal veterinarian)

There was clear disparity in the data between incidents which participants felt needed reporting being those caused by another party, and injuries perceived as self-attributable to be considered less worthy of reporting.

## Discussion

This is the first study to explore how veterinary professionals define work-related injuries, and to compare perception across different veterinary sectors, and job roles.

It was clear that work within the veterinary industry is perceived as being frequently harmful, and that there are social norms around injury expectation which are different across veterinary sectors. For example, needlestick injuries were commonly expected across all practising veterinary job roles, whilst defining an injury as the inability to perform work was primarily seen in production animal and equine veterinarians. These data clarify why there may be differences in reporting of injuries across the veterinary industry, as people are unlikely to report what they consider to be “minor” or “expected” injuries. Nevertheless, within each group there was also considerable variation, which suggests that some professional cultures may not perceive each of these items to be ‘expected’ and therefore many of these harms are avoidable if tackled.

It is notable that accidents were deemed worthy of reporting when caused by an external presence, such as an animal, rather than by staff themselves. This suggests that an element of blame attribution may be perceived as a requirement for reporting, which could be connected with the findings in other industries that events construed as “personal failures” were less worthy of reporting than events with an external cause (14). Injury self-attribution may be deemed as embarrassing, unworthy of sharing, and unlikely to lead to future behaviour change for others. A culture of ‘ownership’ of an injury needs to be embraced, which could facilitate more rapid change in risk mitigation and reporting strategies.

Needlestick injuries are a common incident that was described as not worthy of reporting, likely because they did not fulfil the “injury” criteria: they were perceived as common, minor, self-attributed, and did not cause lasting pain. However, needlestick injuries are harmful, and it was concerning to see that they were considered by some to be an everyday occurrence. This is consistent with previous research which described attitudes of veterinarians to needlestick injuries to be “relatively lax” (5). Needlestick injuries are problematic in terms of frequency and under-reporting in human medicine (19–22). Our study contributes to the explanation for this poor adherence through our finding that these injuries do not meet the criteria commonly applied to identifying an “injury”.

The results of this study indicate that the veterinary industry should take note of the discrepancy of perception of what constitutes work-related injury, in its practices and reporting protocols. This impacts reporting practices and interpretation of injury statistics. Managers within the veterinary industry need to recognise that there is clear differentiation in injury perception between different veterinary sectors and job roles. Yet acknowledging these differences to their staff could widen that divide and imply that certain harms are inevitable in some situations. We suggest that it is important that each collective workplace considers: the culture and attitudes of its employees; what is thought to be an “everyday” occurrence; what is perceived as “acceptable risk”; and how the organisation plans to manage its reporting standards. Managers need to consider what their local workforce consider an acceptable level of risk; what is considered a workplace injury; what is deemed unworthy of reporting; whether staff feel comfortable or have time to reporting safety breaches; and whether staff feel there are useful consequences from reporting. A similar approach is recommended by researchers exploring accidents in hospitals, who recommend exploration of issues by integrating the organisational, individual, and technical factors when examining approaches to risk management (23). Close attention to the human factors within the microculture of each workplace is likely to bring about meaningful change and improve worker safety.

Many veterinary professionals will only report a severe injury, and therefore statistics collected will under-estimate the true prevalence. Industry-wide cultural change needs to occur to support individuals in appreciating the need for reporting and collating of injury data. This has occurred in other industries; a hospital found that an anonymous reporting system increased reporting of incidents by over five times (24), which aligns with other research which supported ‘identification and blame’ as a significant barrier to reporting (25). These findings highlight the need for reporting systems to be presented in ways which match the culture and concerns of the workers in relation to blame attribution and promoting inter-staff harmony.

There are some clear limitations to this study. The number of respondents in equine and production animal practice was relatively low. Secondly, since the survey was disseminated via the research funder, participants might have provided biased answers writing what they felt that their employers “expected”. This is potentially highlighted through the high number of respondents answering that all injuries should be reported. However, the influence was hopefully minimised by reiterating that responses would not be seen or analysed by the funder, and that all responses were anonymous. Finally, the survey methodology led to predominantly short-answer descriptions; while these provide valuable insight, additional information about context and experience would be invaluable in understanding attitudes to reporting and the broader cultural contexts. To explore the differences in attitude versus practice meaningfully, more in-depth observational or ethnographic study is required. Though inductive content analysis is appropriate in a survey with short responses, an absence of mentioning a code does not mean that it lacks importance for those individuals.

## Conclusions

This study provides the first insight into perceptions of work-related injury in veterinary practices across the UK, including a comparison across different job roles. It highlights that defining a work-related injury was a complex and nuanced concept. Each sector of the industry has its own culture of risk and injury expectation. These expectations can impact what people consider to be “everyday” risks, which are not worth reporting. Veterinary practices should explore their own microcultures in relation to accidents, injuries, and reporting practices. Care must be taken interpreting injury statistics as it is likely that under-reporting levels are high, with only the most serious injuries being reported.

This research suggests an individualised approach will lead to targeting appropriate endpoints: some workplaces may need to work on revisiting their ideas around acceptable risk, for example with animal handling; others may be good at minimising work-related harm, but have a complex, or blame-associated reporting system. Considering a work culture, that supports ‘ownership’ of an injury, at a local level is most likely to be successful in creating meaningful change in attitude and behaviour.

## Data availability statement

All data produced in the present study are available upon reasonable request to the authors.

## Ethics approval

The study received ethical approval from the University of Liverpool Veterinary Research Ethics Committee (VREC1256). Consent to participate in the study and for respondents’ data to be published in aggregated or redacted form was sought before respondents partook in the survey. Respondents were reassured that only aggregated or redacted data would be shared with CVS and that no individual would be identifiable.

## Patient consent for publication

Not applicable

## Acknowledgements

We would like to thank all respondents who took part in the study.

## Contributors

Conceptualisation: JT, MW. Funding Acquisition – JT, MW, RJ. Methodology: JT, MW, TF, IS. Formal analysis: JT, TF. Writing original draft – TF, JT. Writing review & editing – All authors

## Rights retention

For the purpose of open access, the author has applied a Creative Commons Attribution (CC BY) licence to any Author Accepted Manuscript version arising from this submission.

## References

1. US Bureau of Labor Statistics. Survey of Occupational Injuries and Illnesses Data. 2023. https://www.bls.gov/iif/nonfatal-injuries-and-illnesses-tables.htm (accessed 1 Dec 2023)

2. US Bureau of Labor Statistics. Incidence rates of nonfatal occupational injuries and illnesses by industry and case types, 2021. 2022. https://www.bls.gov/iif/nonfatal-injuries-and-illnesses-tables/table-1-injury-and-illness-rates-by-industry-2021-national.htm#soii_n17_as_t1.f.1 (accessed 1 Dec 2023)

3. Parkin TDH, Brown J, Macdonald EB. Occupational risks of working with horses: A questionnaire survey of equine veterinary surgeons. Equine Vet Educ 2018; Apr 29;30(4):200–5. doi:10.1111/eve.12891

4. Tulloch JSP, Fleming KM, Pinchbeck G, et al. Audit of animal-related injuries at UK veterinary schools between 2009 and 2018. Vet Rec 2023; Oct 7;193(7). doi: 10.1002/vetr.3171

5. Weese JS, Jack DC. Needlestick injuries in veterinary medicine. Can Vet J 2008;49(8):780–4.

6. Stout N, Linn H. Occupational injury prevention research: progress and priorities. Inj Prev 2008;8:iv9–14. doi: 10.1136/ip.8.suppl_4.iv9

7. Vandeskog B. The Legitimacy of Safety Management Systems in the Minds of Norwegian Seafarers. TransNav, Int J Mar Navig Saf Sea Transp 2015;9(1):101–6. doi: 10.12716/1001.09.01.12

8. CDC. Definitions for WISQARS™ Nonfatal. 2018. https://www.cdc.gov/injury/wisqars/nonfatal_help/definitions.html#:~:text=Nonfatal xInjury Reports-,5.1 Definition of Nonfatal Injury,be unintentional or violence-related. (accessed 1 Dec 2023)

9. Health and Safety Executive. Key definitions. https://www.hse.gov.uk/riddor/key-definitions.htm (accessed 1 Dec 2023)

10. Hallowell M. Safety risk perception in construction companies in the Pacific Northwest of the USA. Constr Manag Econ 2010; Apr;28(4):403–13. doi: 10.1080/01446191003587752

11. Paudel S, Subedi M, Brangan E, et al. 4C.001 Conceptualising ‘injury’ in Nepal: Building shared understandings as a foundation for engagement. Inj Prev 2021;Mar;27:A34.3–A35. doi:10.1136/injuryprev-2021-safety.104

12. Edmund NNK, Suxia L, Ebenezer L, et al. Emotional intelligence as a conduit for improved occupational health safety environment in the oil and gas sector. Sci Rep 2023; Nov 11;13(1):19698. doi: 10.1038/s41598-023-46886-3

13. Kyung M, Lee S-J, Dancu C, et al. Underreporting of workers’ injuries or illnesses and contributing factors: a systematic review. BMC Public Health 2023; Mar 24;23(1):558. doi: 10.1186/s12889-023-15487-0

14. Bhattacharya S. Sociological factors influencing the practice of incident reporting: the case of the shipping industry. Empl Relations 2011; Nov 11;34(1):4–21. doi: 10.1108/01425451211183237

15. Vears DF, Gillam L. Inductive content analysis: A guide for beginning qualitative researchers. Focus Heal Prof Educ A Multi-Professional J 2022; Mar 31;23(1):111–27. doi: 10.11157/fohpe.v23i1.544

16. Krippendorff K. Content Analysis: An Introduction to Its Methodology [Internet]. 2455 Teller Road, Thousand Oaks California 91320: SAGE Publications 2019.

17. Robinson D, Edwards M, Mason B,et al. The 2019 survey of the veterinary profession [online]. 2019. https://www.rcvs.org.uk/news-and-views/publications/the-2019-survey-of-the-veterinary-profession/?destination=%2Fnews-and-views%2Fpublications%2F%3Ffilter-keyword%3D%26filter-type%3D%26filter-month%3D%26filter-year%3D2020%26p%3D2&&&type=rfst&set=true#cookie-widget (accessed 1 Dec 2023)

18. Robinson D, Edwards M, Akehurst G, et al. The 2019 Survey of the Veterinary Nurse Profession [online]. 2019. https://www.rcvs.org.uk/news-and-views/publications/the-2019-survey-of-the-veterinary-nursing-profession/ (accessed 1 Dec 2023)

19. Cooke CE, Stephens JM. Clinical, economic, and humanistic burden of needlestick injuries in healthcare workers. Med Devices Evid Res 2017; Sep;10:225–35. doi: 10.2147/MDER.S140846

20. Callan RS, Caughman F, Budd ML. Injury Reports in a Dental School: A Two-Year Overview. J Dent Educ 2006; Oct; 70(10):1089–97. doi: 10.1002/j.0022-0337.2006.70.10.tb04182.x

21. Behzadmehr R, Balouchi A, Hesaraki M, et al. Prevalence and causes of unreported needle stick injuries among health care workers: a systematic review and meta-analysis. Rev Environ Health 2023; Mar 28;38(1):111–23. doi: 10.1515/reveh-2021-0148/html

22. Motaarefi H. Factors Associated with Needlestick Injuries in Health Care Occupations: A Systematic Review. J Clin Diag Res 2016; 10(8):IE01–IE04. doi: 10.7860/JCDR/2016/17973/8221

23. Barach P. Reporting and preventing medical mishaps: lessons from non-medical near miss reporting systems. BMJ 2000; Mar 18;320(7237):759–63. doi: 10.1136/bmj.320.7237.759

24. Stump LS. Re-engineering the medication error-reporting process: Removing the blame and improving the system. Am J Heal Pharm 2000; Dec 1;57:S10–7. doi: 10.1093/ajhp/57.suppl_4.S10

25. Carroll JS. Organizational Learning Activities in High-hazard Industries: The Logics Underlying Self-Analysis. J Manag Stud 1998; Nov 16;35(6):699–717. doi: 10.1111/1467-6486.00116

